# Short- and long-term causes of West Nile virus risk in Europe: a spatiotemporal model accounting for under-reporting

**DOI:** 10.64898/2026.08.20.26360902

**Authors:** Jonathan Bastard, Charles Assaad, Renaud Marti, Annelise Tran, Raphaëlle Métras, Benoit Durand

## Abstract

Models that provide risk maps for zoonoses often lack (i) a spatiotemporal autocorrelation component, yet crucial in understanding the spread of infectious diseases, (ii) accounting for heterogeneity in case reporting, and (iii) a causal framework for explanatory variables. Here, we addressed these limitations with a model system, West Nile virus, a vector-borne pathogen transmitted in a bird reservoir, and affecting humans and horses. We built a spatiotemporal occupancy model and fitted it to notified (human and horse) case data. Based on a directed acyclic graph, we estimated the causal effects of conjectural weather variables (i.e. changing in the short-term) *vs*. structural variables (i.e. changing in the long-term) on WNV circulation in the bird reservoir, besides assessing variables associated with case reporting. By computing population attributable fractions, we found the contribution of conjectural weather variables to WNV outbreaks in Europe to be globally higher than the structure of the bird community.

## Introduction

Understanding and predicting the occurrence of pathogens is often hampered by their imperfect and heterogeneous detection. This is particularly the case for zoonotic vector-borne viruses because (i) their circulation in the animal reservoir may be little known, (ii) infection in humans may cause no or unspecific symptoms (e.g. flu-like symptoms), and (iii) their detection relies on surveillance systems that may differ across species and regions ^1^.

As a model system, West Nile virus (WNV) is a zoonotic orthoflavivirus transmitted between reservoir birds and *Culex* mosquitoes. It also affects mammals, including humans, considered as “dead-end hosts” because they cannot infect biting mosquitoes ^2^. However, human-to-human transmission is still possible through blood donations and organ transplantations ^3^. In humans, while asymptomatic infections are most frequent, the virus can cause dengue-like symptoms and, more rarely, neurological disease that may lead to hospitalization and death ^2^. This is why it is important to identify geographical patterns of WNV circulation and its drivers, even though it may go unnoticed.

In Europe, between 100 and 2000 WNV human cases are notified annually to the European Centre for Disease Prevention and Control (ECDC) ^4^. Animal cases, mostly horses and wild birds, are also frequently reported to the World Organisation for Animal Health ^5^. The pathogen has been long present around the Mediterranean Basin and in Central and Eastern Europe, but its geographical range has recently expanded in Europe, with Germany reporting cases since 2018, the Netherlands since 2020, western and northern regions of France since 2022, and Baltic countries since 2024 ^4^.

In a given region, the risk of WNV circulation depends on mechanisms changing across various timespans, from the landcover type affecting what host species are present to quickly changing weather conditions impacting mosquito vector density. Hence, we can partition WNV risk into structural (quasistatic or long-term) and conjectural (dynamic or short-term) components ^6^.

Previous studies modelled the occurrence of WNV across European regions ^7–15^ but exhibited the following limitations. First, few accounted for spatio-temporal autocorrelation as part of the model structure, despite its potentially strong impact on cases spatial and temporal distribution. Not accounting for this autocorrelation by treating regions as independent epidemiological units might then bias estimates of predictors’ effects. Second, most of them relied on human cases as reported by ECDC surveillance system, and more rarely on animal cases ^7,12^, whereas WNV has been shown to be able to spread silently without causing reported symptomatic human or animal cases ^16^. Third, previously published models were maximized with regards to their predictive capacities, at the possible expense of their explanatory capacities. Indeed, Yates and colleagues ^17^ argue that the objective of maximizing a model’s capacity to predict new data may not be always compatible with causal inference focused on parameter estimation.

Here, we developed a spatio-temporal occupancy model with two components quantifying WNV circulation in the bird reservoir on the one hand, and the subsequent reporting of cases on the other hand. By fitting it to human and animal cases notified from the European Union between 2010 and 2024, we assessed WNV infection risk, disentangling the risk attributable to long-term (structural component of the risk) and short-term (conjectural component of the risk) predictors, and accounting for the spatial and temporal structure of the data, as well as the heterogeneity in cases detection across countries.

## Results

### Data

By collating data from ECDC and the World Animal Health Information System (WAHIS), we gathered reports of WNV human and equine cases notified in the 27 European Union countries for the period 2010-2024 ^4,5^. Figure 1 shows that their spatial distribution was heterogenous, with most affected areas in southern, central and eastern parts of Europe. Overall, the number of NUTS administrative regions reporting at least one human case (246 regions out of 727) was higher than those reporting at least one equine case (106 regions). Within-year, the peak of reported human cases was in August, and a month later for reported equine cases (Figure 1), and 98% of NUTS-months with cases were between June and October.

**Figure 1.**
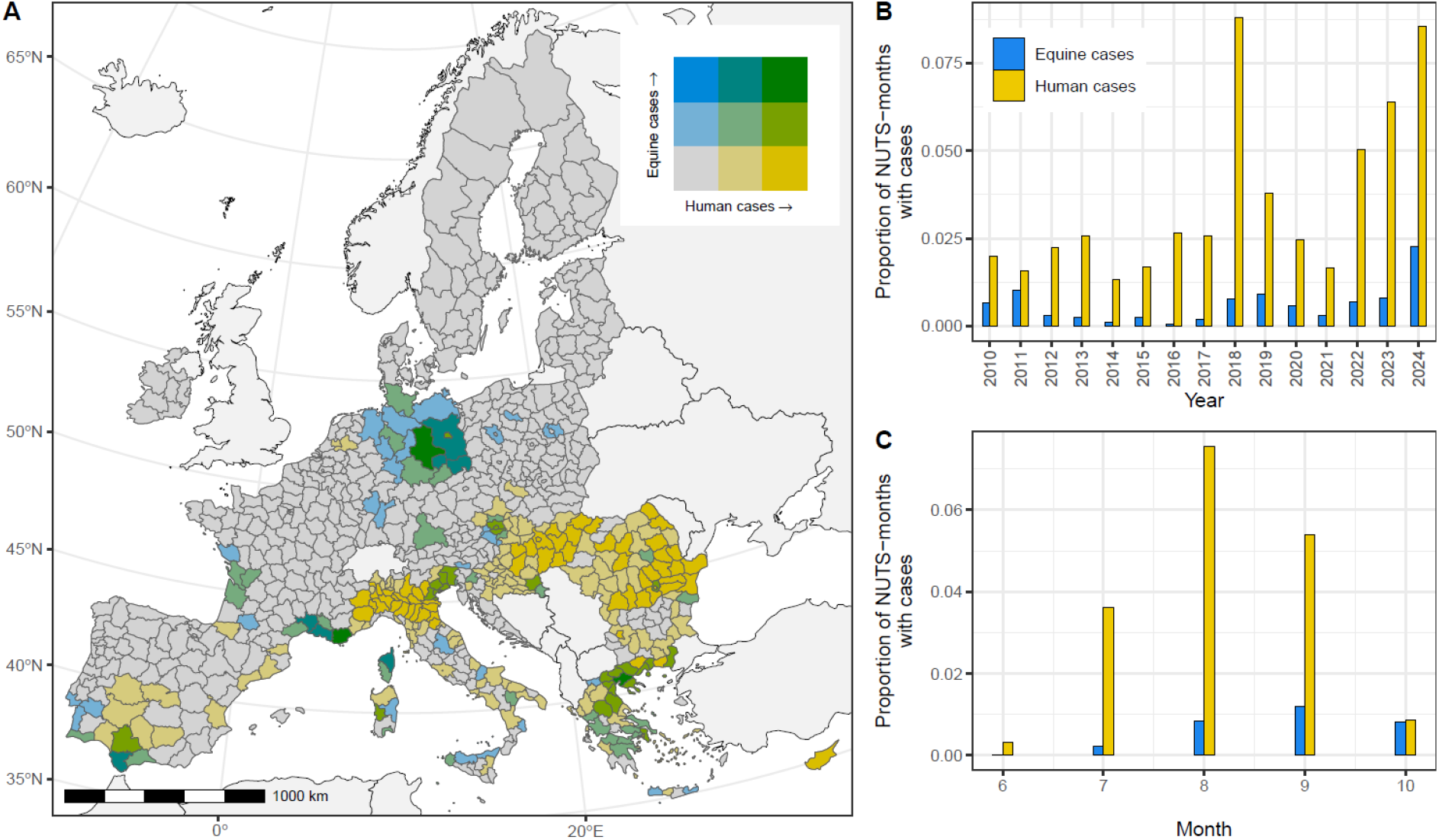
WNV cases data collated and analyzed in the study. Proportion of NUTS-months with human or equine cases in the European Union between 2010 and 2024 (June to October, i.e. the warm season). NUTS regions are administrative regions of the European Union. We used the NUTS3 level, excepted in four countries (Belgium, Germany, Malta and Netherlands) where all data was aggregated at the NUTS2 level. In panel A, we combine three categories for notified human cases and three categories for notified equine cases: 0%, between >0 and 10%, and >10% of months with cases (the denominator is the total number of June-to-October months during the study period, i.e. 75 months). In the right panels, we display the proportions of NUTS-months with WNV human and equine cases by year (panel B) and by month of the year (panel C) included in the analysis. The denominator is 3,635 NUTS-months in panel B (5 warm-season months x 727 NUTS), and 10,905 NUTS-months in panel C (15 years x 727 NUTS).

### Model

To investigate the spatial and temporal patterns of WNV infections, we built a spatio-temporal occupancy model of the virus in Europe, with both a “Circulation” and a “Reporting” component (Figure 2). The “Circulation” component had the same structure in the “Equine” and “Human” models, and aimed at assessing the impact of structural (i.e. changing on the long-term) *vs*. conjectural factors (i.e. changing on the short-term such as weather variables) affecting viral circulation in the bird reservoir, to which humans and equids are then exposed. The causal structure of these “Circulation” variables was determined using a Directed Acyclic Graph (DAG) (Supplementary Figure S1). The “Reporting” component was species-specific, and included demographic and socio-economic variables.

**Figure 2.**
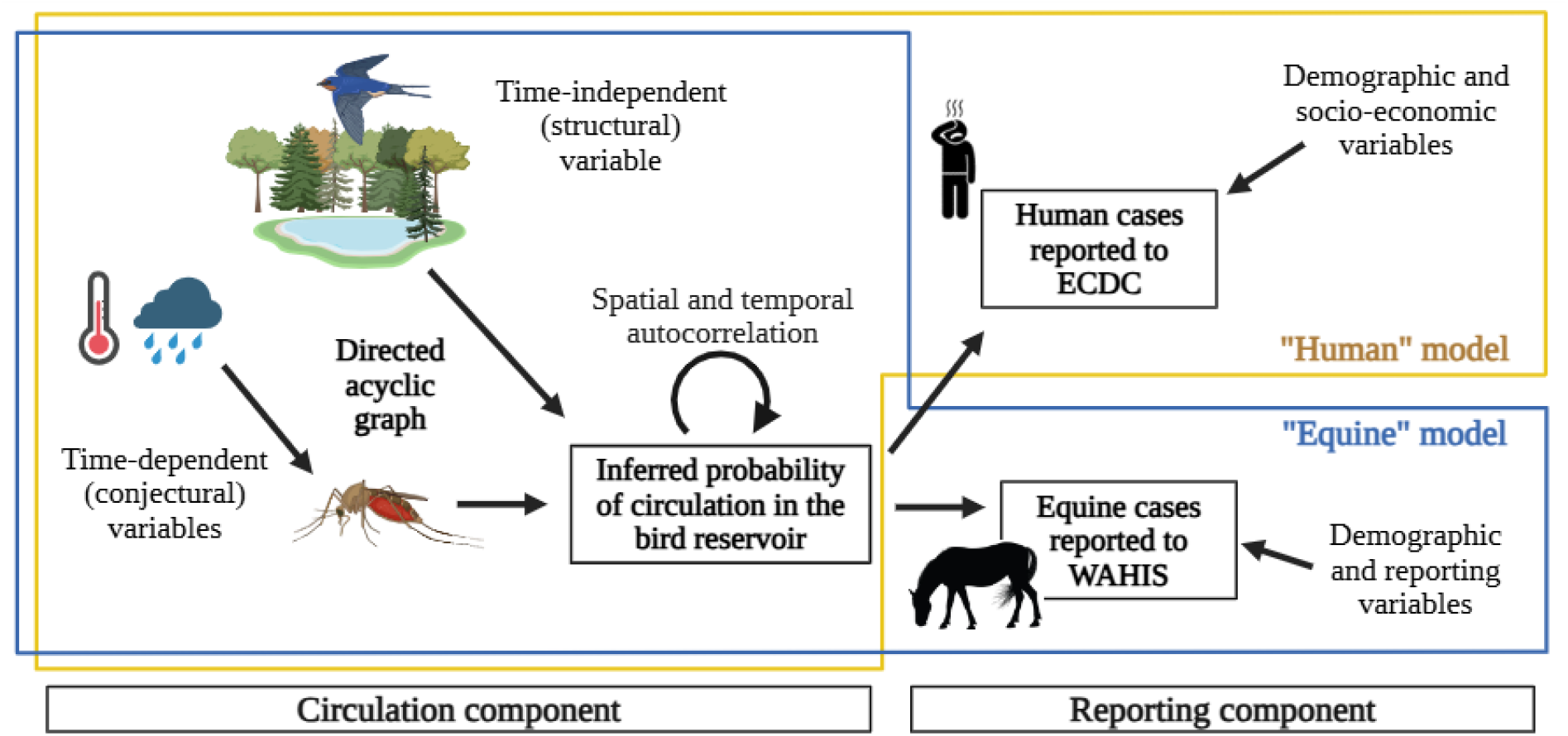
Diagram of West Nile virus (WNV) spatio-temporal occupancy model. In the “Virus circulation” component, both structural (time-independent) and conjectural (time-dependent) variables are related through a directed acyclic graph (DAG) where the outcome is the (unobserved) probability of WNV circulation in the bird reservoir in a given NUTS region for a given month. Spatial and temporal autocorrelation are accounted for with Besag-York-Mollie 2 (BYM2) and Randow walk processes. The “Case reporting” component allows to fit the model to the human or equine case data (“Human” and “Equine” model respectively), using demographic and socio-economic variables and the probability of circulation from the “Circulation” component. Figure created with BioRender.com.

Model results showed that, between 2010 and 2024, WNV circulated in reservoir birds mostly in Eastern Germany and other Central Europe areas, Greece, Southern Spain, Southern France and, in the “Human” model specifically, Northern Italy, Romania and Hungary (Figure 3). Regarding these latter areas, the discrepancies between the “Human” and “Equine” model predictions were consistent with differences between the observed spatial distribution of reported human *vs*. equine cases (Figure 3).

**Figure 3.**
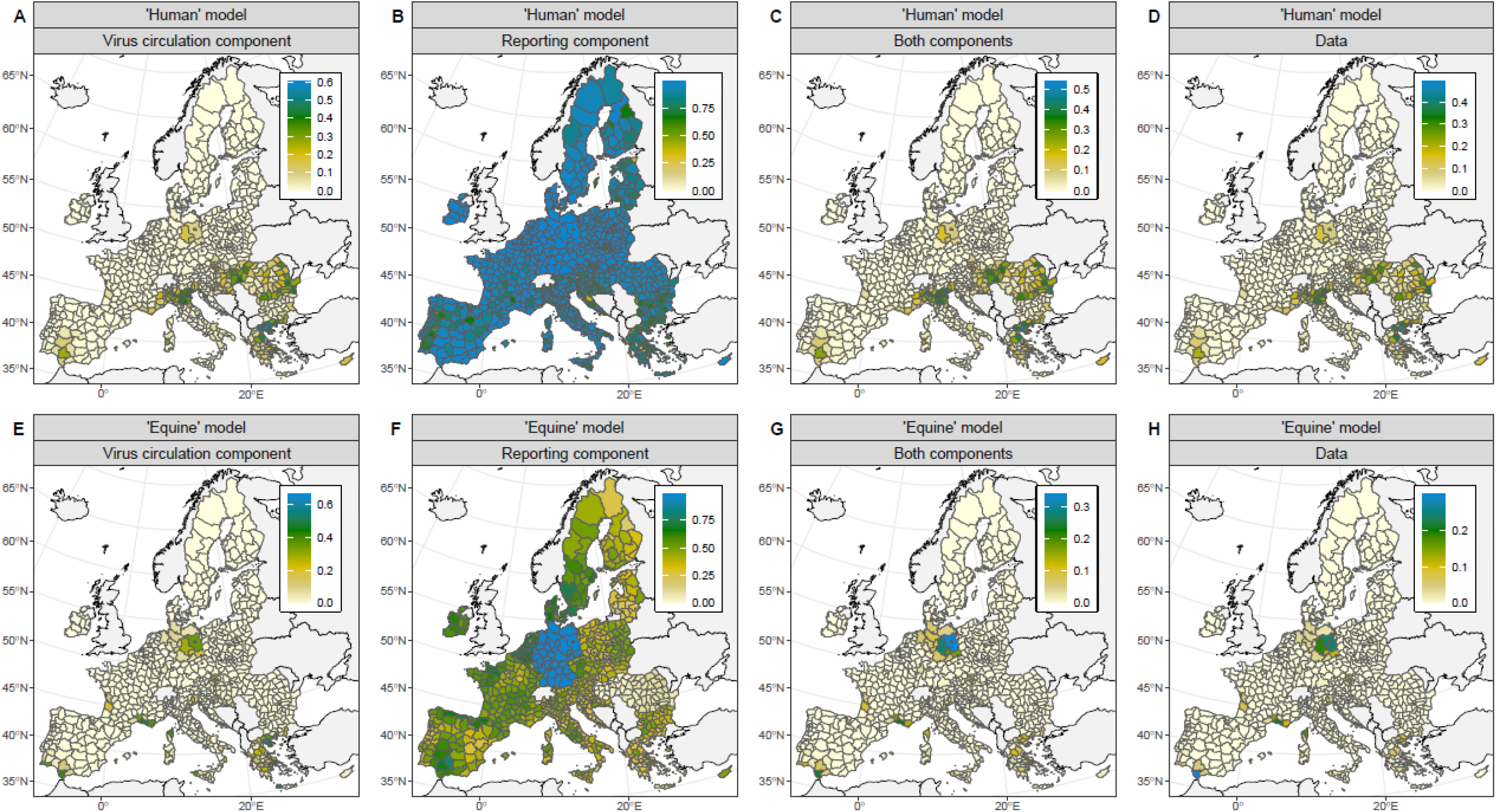
Model predictions and goodness of fit. Across months between 2010 and 2024 (June to October), average probabilities of WNV circulation in the bird reservoir (panels A and E), of WNV case reporting given circulation in the bird reservoir (panels B and F) and of overall WNV case reporting (panels C and G), predicted by the “Human” (top panels) and “Equine” (bottom panels) models, and observed proportion of months with reported human (panel D) and equine (panel H) cases. Proportions are computed by removing months with missing data in each NUTS region.

During cross-validation of the occupancy models, the pooled deviance-based marginal R^2^ was 0.03 for the “Equine” model, and 0.23 for the “Human” model, showing the predictive value added by the models’ fixed effects on top of random (including spatial and temporal) effects. Moreover, the NUTS and months with a predicted detection of WNV in Europe were consistent with both the fitting dataset (reported human and equine case data, AUC of respectively 0.97 and 0.98) and the validation dataset (wild bird outbreaks reported to WAHIS, AUC of respectively 0.87 and 0.93) (Figure 3 and Supplementary Figures S2 and S3).

### Effects of structural and conjectural variables on WNV circulation

Under our DAG assumptions, the coefficients for some variables (namely the Bird Risk Index, BRI, and the anomalies in temperature and in the Modified Normalized Difference Water Index, AnomTemp^m^ and AnomMNDWI^m^) could be considered as causal effects on the probability of WNV circulation in the bird reservoir at the month and NUTS region scale (Supplementary Figure S1). On the contrary, coefficients for the other variables could only be considered as associations given the model’s adjustment set. For instance, the coefficient estimated for AbsTemp^m^ was the effect on WNV circulation passing neither through AnomTemp^m^ nor through AnomMNDWI^m^, therefore it could not be considered as a total causal effect (Supplementary Figure S1).

In the “Equine” model, we estimated a positive causal effect of AnomTemp^m^ on WNV circulation (estimate and 95% credible interval of 0.45 [0.23; 0.80]), and associations between multiple other conjectural weather variables (namely AnomTemp^m-1^, AbsTemp^m^, AbsTemp^m-1^, AbsTemp^spring^, AnomMNDWI^m-1^ and AnomMNDWI^spring^) and this outcome (Figure 4). The estimated causal effect of the BRI, a structural variable quantifying the composition of the bird reservoir population ^18^ was not different from 0 in this model (0.07 [-0.41; 0.54]) (Figure 4). In Supplementary Table S1, we expressed these effects in terms of odds-ratios relative to unscaled variables. For example, in a given NUTS, a monthly temperature increased by one standard deviation as compared to the 2002-2024 period caused an odds-ratio of 1.61 [1.28; 2.32] in the “Equine” model (Supplementary Table S1).

**Figure 4.**
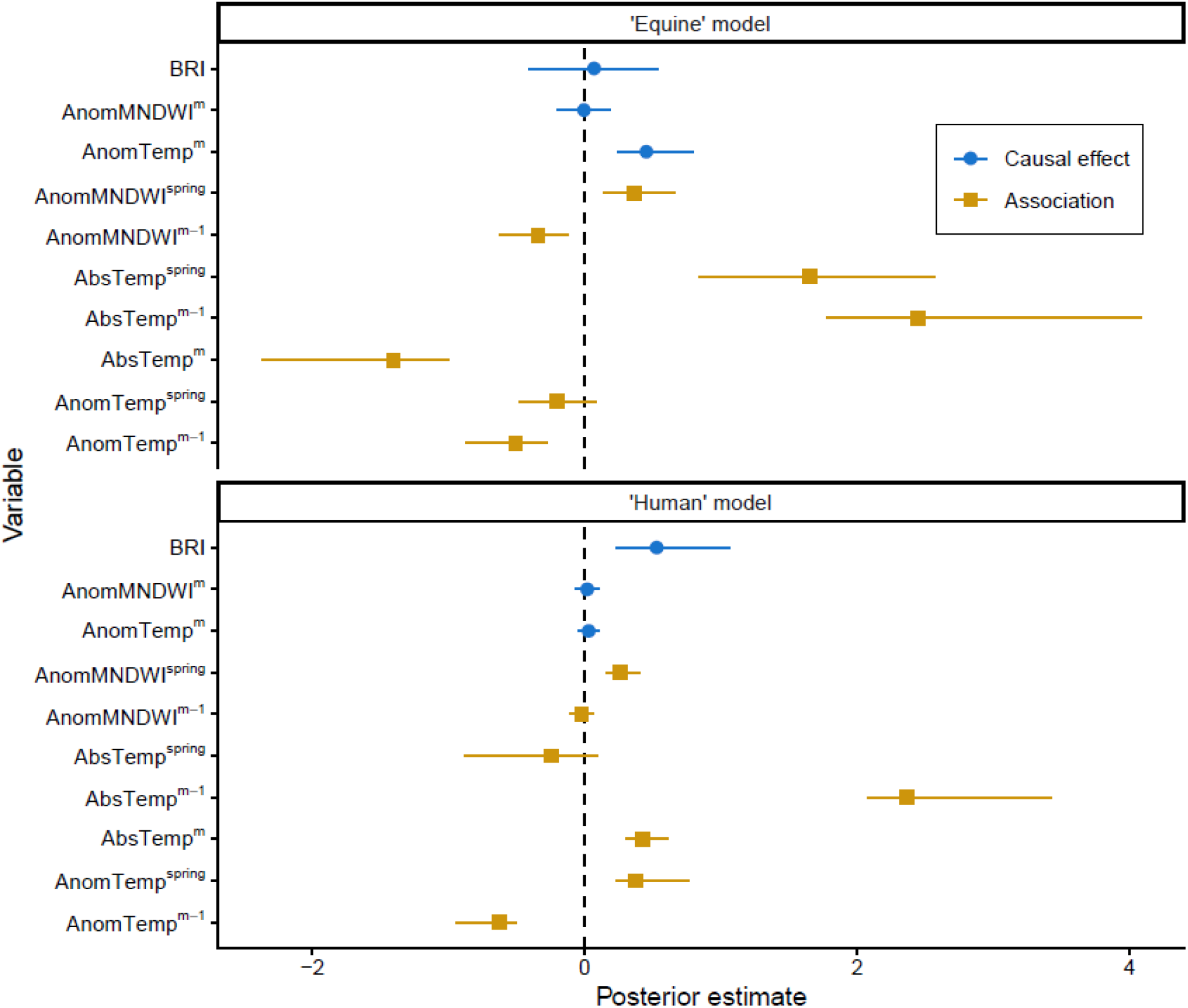
Variable coefficients of the “Circulation” component. In both “Equine” and “Human” models, under our DAG assumptions, the coefficients for some variables can be considered as causal effects on the probability of WNV circulation in the bird reservoir on month m at the NUTS region scale (blue circles), while others are only associations (brown squares). Points and intervals are respectively the posterior median and 95% credible interval.

In the “Human” model, we also found associations between several weather variables (AnomTemp^m-1^, AnomTemp^spring^, AbsTemp^m^, AbsTemp^m-1^, AnomMNDWI^m-1^ and AnomMNDWI^spring^) and WNV circulation, although no conjectural causal effect differed from 0 (Figure 4). However, we estimated a positive causal effect of the structural BRI in this model (0.45 [0.23; 0.80]). This corresponded to an increase of the BRI by one interquartile range causing an odds-ratio of 1.85 [1.30; 3.44] (Supplementary Table S1).

When we analyzed the sensitivity of causal effect estimates to different sets of adjustment variables, we inferred approximately the same values as in the main analysis, supporting the robustness of our results and of the DAG assumptions (Supplementary Figure S4).

### Population attributable fractions

Using model estimates, we computed population attributable fractions (PAF) ^19–21^ to compare the relative impacts of the structural *vs*. conjectural variables on the probability of WNV circulation at the month and regional level. The global PAF for the group of conjectural (weather) variables was 2.5 times that of the one structural variable (the BRI) in the “Human” model (99.6% *vs*. 39.8%), and 15.5 times in the “Equine” model (99.9% *vs*. 6.5%), suggesting a strong effect of short-term weather determinants on the risk of WNV circulation in Europe.

When analyzed by NUTS region, the structural PAF was heterogeneously distributed across Europe in both models, with higher values on the Eastern part of the continent, as expected by the spatial distribution of the BRI ^18^ (Figures 5 and 6). In the “Human” model, the conjectural PAF was higher in the South of Europe and in the Balkans (Figure 5), suggesting favorable weather conditions for WNV circulation during the warm season in these areas. On the contrary, the more homogeneous spatial distribution of the conjectural PAF from the “Equine” model (Figure 6), excepted some regions in the Alps and Scandinavia, may reflect the widespread notification of equine cases in Europe.

**Figure 5.**
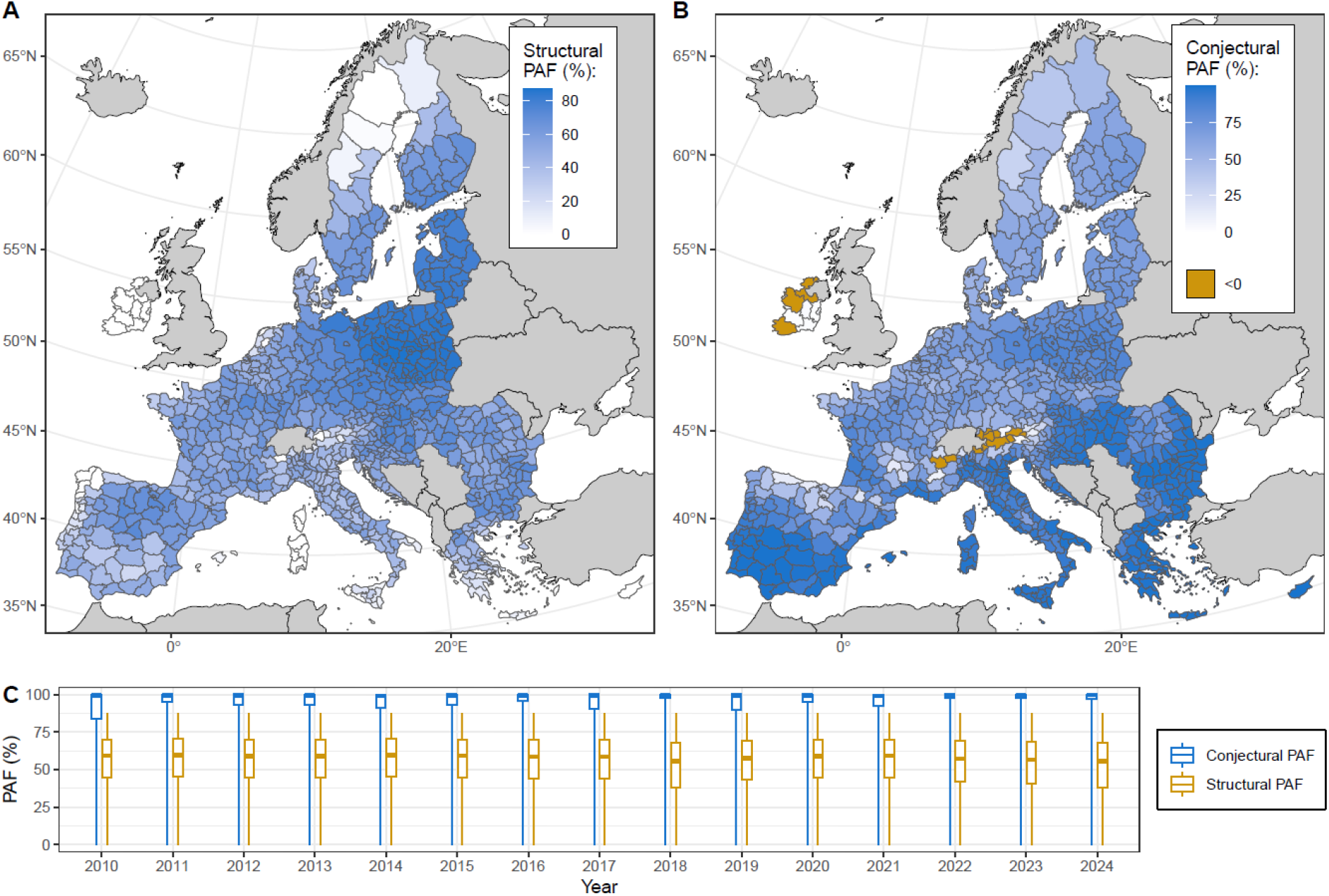
Population attributable fractions (PAF) in the “Human” model. Structural and conjectural PAF were computed for each NUTS-month using the same method than the global PAF. From the “Human” model, panels A and B show respectively the structural and conjectural PAF averaged in each NUTS region, while panel C shows the distribution of both PAF for each year (the box plot represents the minimum, 25^th^ percentile, median, 75^th^ percentile and maximum). Due to the computation method, the conjectural PAF for some NUTS-months can be <0. For visualization, panel C was bounded below by 0.

**Figure 6.**
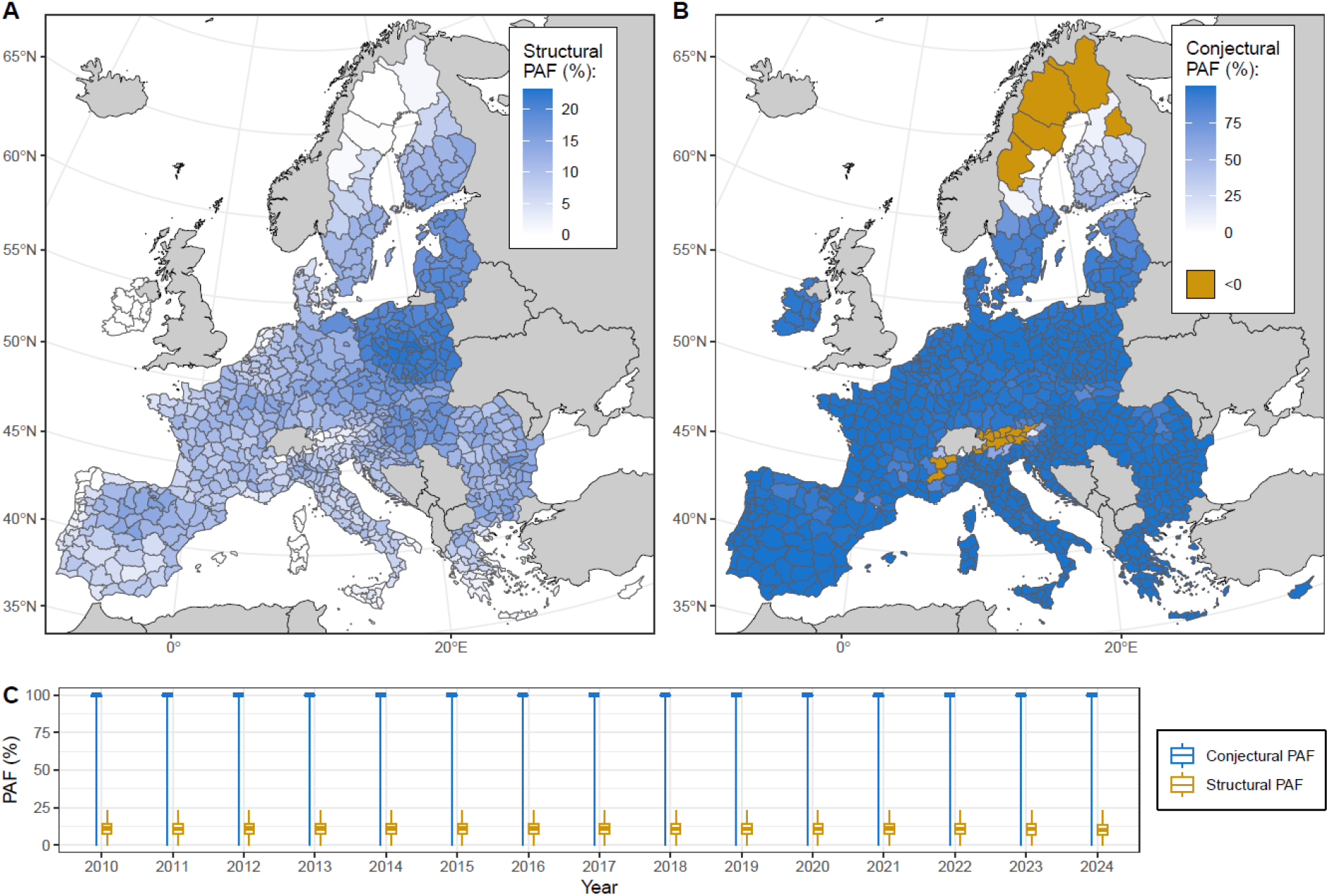
Population attributable fractions (PAF) in the “Equine” model. Structural and conjectural PAF were computed for each NUTS-month using the same method than the global PAF. From the “Equine” model, panels A and B show respectively the structural and conjectural PAF averaged in each NUTS region, while panel C shows the distribution of both PAF for each year (the box plot represents the minimum, 25^th^ percentile, median, 75^th^ percentile and maximum). Due to the computation method, the conjectural PAF for some NUTS-months can be <0. For visualization, panel C was bounded below by 0.

Although always higher than the structural PAF, the year-aggregated conjectural PAF presented some annual variations with, for example, weather conditions more favorable to WNV circulation in 2018 and 2022 to 2024 in the “Human” model (Figure 5).

### Variables associated with WNV reporting

In the occupancy model, different variables affected the probability of WNV circulation in the bird reservoir, and the probability of subsequent detection (reporting) following reservoir circulation in a given NUTS and month (Figure 1). The latter was estimated to a median of 0.93 for human cases, and of 0.39 for equine cases, although with spatial heterogeneity (Figure 3). The human log-population (LogHumPop) and being classified by Eurostat as an urban NUTS region (UrbClass) were found to be respectively positively and negatively associated with the reporting of human cases (1.25 [0.84; 1.67] and -2.01 [-3.20; -0.83]) (Table 1). As expected, the equine log-population (LogEqPop) and being classified as a country that may over-report WNV equine cases as compared to other European countries (CountryOverReport, namely Germany) were positively associated with the reporting of equine cases (0.88 [0.54; 1.22] and 2.99 [1.34; 4.72]), while being classified as a country that may under-report WNV equine cases (CountryUnderReport, namely Hungary and Romania) was negatively associated (-2.66 [-4.34; -0.97]) (Table 1).

**Table 1.**
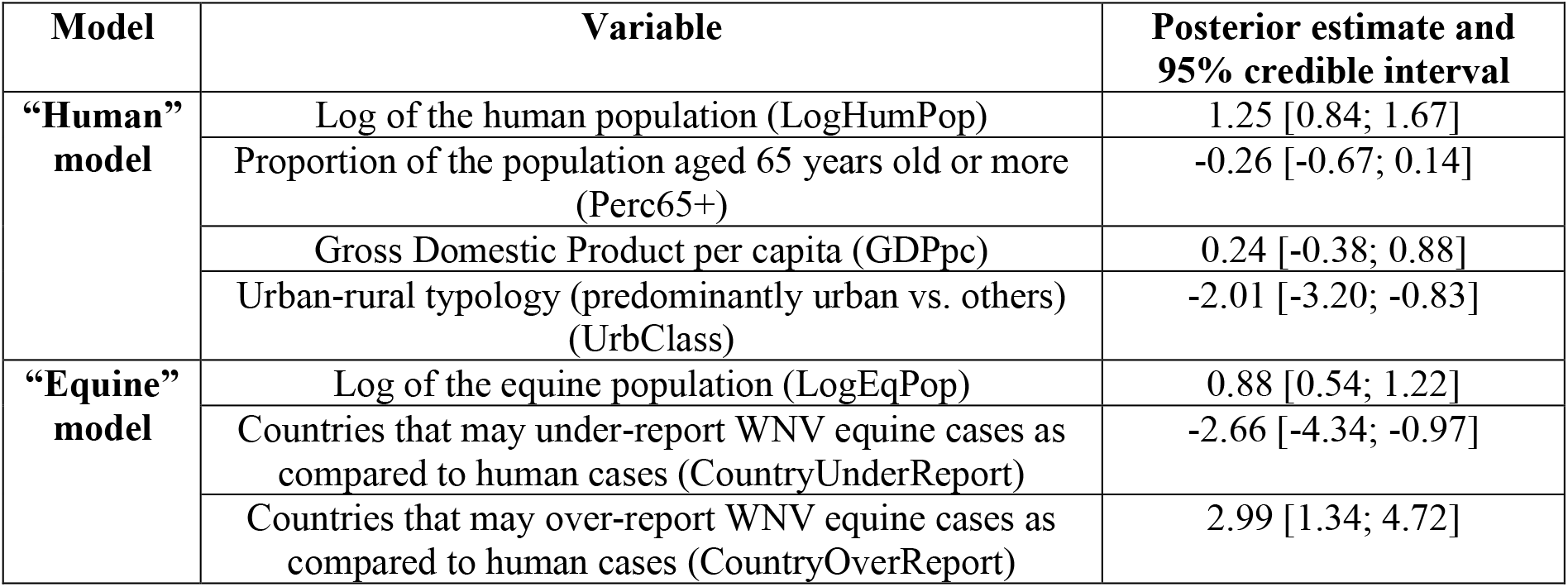
Variable coefficients of the “Reporting” component. Variables included in the “Reporting” component depend on the model. Values and intervals are respectively the posterior median and 95% credible interval.

| Model | Variable | Posterior estimate and 95% credible interval |
| --- | --- | --- |
| “Human” model | Log of the human population (LogHumPop) | 1.25 [0.84; 1.67] |
|  | Proportion of the population aged 65 years old or more (Perc65+) | -0.26 [-0.67; 0.14] |
|  | Gross Domestic Product per capita (GDPpc) | 0.24 [-0.38; 0.88] |
|  | Urban-rural typology (predominantly urban vs. others) (UrbClass) | -2.01 [-3.20; -0.83] |
| “Equine” model | Log of the equine population (LogEqPop) | 0.88 [0.54; 1.22] |
|  | Countries that may under-report WNV equine cases as compared to human cases (CountryUnderReport) | -2.66 [-4.34; -0.97] |
|  | Countries that may over-report WNV equine cases as compared to human cases (CountryOverReport) | 2.99 [1.34; 4.72] |

## Discussion

We built a spatiotemporal model to explain West Nile virus occurrence across Europe, accounting for the causal effects of time-independent (structural or long-term) variables, time-dependent (conjectural or short-term) weather variables, and a heterogeneous reporting of human and equine cases.

Among short-term weather drivers, we found a positive causal effect of temperature anomaly in summer months on WNV circulation in the bird reservoir on the same months, in the “Equine” model, although it was not the case for MNDWI anomaly. This was consistent with previous studies that tended to show positive associations between temperature and WNV infections ^22,23^, despite difficulties to compare different outcomes (e.g. mosquito infection prevalence *vs*. human cases) and spatial scales and climates (e.g. Mediterranean climate or wetland ecosystem over a few hectares *vs*. the diversity of climates over the entire Europe). Our result might be due to temperature anomalies allowing to get closer to WNV optimal transmission temperature as was previously derived by combining pathogen- and mosquito-traits ^24,25^. In both of our models, the combination of all conjectural (short-term) weather factors explained WNV circulation risk more substantially than its structural (long-term) counterpart at the global scale, despite spatial heterogeneities at the NUTS region scale.

However, our study does not preclude a longer-term effect of climatic variables on WNV occurrence. Indeed, our findings suggest a quite strong causal impact of the Bird Risk Index in the “Human” model, i.e. an effect of the bird community composition, which itself depends largely on land cover and on long-term climatic trends ^26^. Moreover, previous publications did show a relationship between climate change and growing WNV outbreaks in Europe ^9,10^, for instance through warmer springs that lead to longer mosquito seasons and thus increased viral transmission ^27^. Here, the causal framework we considered in our model did not allow to isolate the causal effect of spring climatic conditions on WNV outbreaks from other weather drivers, although we did find statistical associations between spring absolute temperature (in the “Equine” model) or spring temperature anomaly (in the “Human” model) and WNV circulation.

Indeed, while previous publications aimed to predict WNV case reports across Europe ^8–11,13,15^, our objective was rather to propose an explanatory framework for the pathogen’s occurrence and detection, using a Directed Acyclic Graph ^28^. This allowed to determine what variables to include or not in the model, and what coefficients could be interpreted as causal effects *vs*. as associations only. In the future, expanding the usage of DAG within the field of vector-borne disease epidemiology may help understanding such complex, multifactorial, eco-epidemiological systems ^29^. However, a limitation that follows this point is that our findings are somewhat conditioned by the causal structure we assumed between variables, and we might not have accounted for some confusion factors or for some relationships between nodes of the DAG. Nevertheless, our approach precisely allows to build on this work, to propose alternative variable networks, and even possibly to orient data collection to test new research hypotheses.

Furthermore, modelling spatial and temporal autocorrelation allowed us to account for the geographical expansion of the virus over time, and therefore to avoid the assumption, implicitly made in many previously published models, that WNV is already present everywhere it would be suitable to. Indeed, the pathogen has been recently expanding in Europe with reports in Germany since 2018, in the Netherlands since 2020, in western and northern areas of France since 2022, and in Baltic countries since 2024 ^4^. The hypothesized mechanisms for these long-distance spreads include movements of infected birds or vector dispersal with the winds ^14,30–32^ although, once introduced in a geographical area, the capacity of the virus to maintain an enzootic cycle depends on the introduction timing, on the mosquito population and on the bird community structure that, here, we modelled as the BRI. The Random Walk process we included allowed to represent the annual variations in WNV circulation, a well-known phenomenon with some years (e.g. 2018) showing more WNV cases than usual ^4^. However, it could not mimic more complex mechanisms that contribute to drive viral dynamics, such as high seroprevalences in horses limiting the extent of outbreaks in following years ^16^.

The dead-end host species used to fit the model, humans or equids, affected causal effect estimates. This may reflect that the infectious hazard, i.e. the probability of WNV circulation inferred by the model, or the exposure to this hazard, do not have the same meaning in the “Equine” than in the “Human” model. First, our findings should be interpreted with regards to the study’s scale, i.e. European NUTS regions, and our analysis is naïve to mechanisms occurring at smaller scales. Yet, humans and equids do not have the same habitat within a given region. Hence, the within-region spatial distribution of both animal and human populations may matter such as, for example, the proximity of stables *vs*. human dwellings to mosquito breeding sites and to geographical areas of possible importance for WNV bird reservoir communities ^33,34^. Likewise, within-region mobility patterns related to work or leisure activities may affect human and equine exposure to infected mosquitoes ^35^. Second, WNV outbreaks are characterized by complex interactions between multiple species (or subspecies) of mosquito vectors with heterogenous feeding patterns, and multiple species of avian and non-avian hosts with heterogeneous transmission competence ^36,37^. Therefore, the exposure of equids *vs*. humans to different mosquito species that previously bit different bird communities may explain variations between “Human” and “Equine” model results. This might be partially reflected by the fact that, in our data and model predictions, the peak in human infections was a month prior to the peak in horse infections. Third, these discrepancies might also be related to biases in the reporting data, although we did account for possible disparities between equine and human WNV reporting fractions as part of the model structure itself.

Indeed, the occupancy modelling framework allowed to differentiate the factors affecting the reporting of (human and equine) cases, given WNV circulation in the bird reservoir, from the factors precisely affecting that circulation ^38,39^. For both species, the log-population in a NUTS region was positively associated with the reporting of cases in that region, which was an expected effect of increasing the exposed population. Given WNV circulation in the bird reservoir, we found the subsequent reporting of human cases to be lower in urban NUTS regions. A possible explanation could be a lower average infection prevalence of *Culex* vectors in urban than in rural settings, because they might feed more on humans, who are dead-end hosts, in an environment with higher human/bird host ratio. This point is still debated ^40^, with some studies showing an increased proportion of blood meals taken in humans as compared to birds in urban areas ^41,42^, and some others that do not ^43,44^. Moreover, we did not observe any association between the country-level GDP per capita and the reporting of human cases, even though a previous publication reported a positive correlation between this macroeconomic index and a country’s capacity to detect global health threats ^45^. Similarly, despite WNV leading to more severe forms and more hospitalizations in older people ^2^, we did not find the proportion of the population above 65 years old to affect human case reporting at the studied scale.

To conclude, our modelling framework allowed to disentangle drivers of WNV spread and detection across Europe. A combination of short-term weather drivers was a key causal factor of WNV outbreaks, although their relative importance as compared to longer term drivers depended on the dead-end host species (human or equid) considered, and on the spatial area considered within Europe.

## Material and Methods

### Data

We analyzed data on WNV notified cases in either the human or equine compartments. On the one hand, for all European Union countries, we obtained from the ECDC the number of human cases reported by month between 2010 and 2024, and by administrative region of the European Union, named NUTS3 regions (Figure 1) ^4^. On the other hand, we extracted WNV event notifications in equids available from WAHIS for the same time period ^5^. Event information includes the species affected, time and geographic coordinates, from which we retrieved the NUTS region. However, equine data from WAHIS were missing for some NUTS and months (Supplementary Method S1 and Supplementary Figure S5).

In four countries (Belgium, Germany, Malta and Netherlands), all data were aggregated at the NUTS2 level instead of NUTS3, because NUTS2 surface area in these countries was closer to the European NUTS3 surface area average. We restricted the analysis to the period from June to October that represented 98% of all NUTS-months with notified cases.

First, we used time-varying weather variables that we named conjectural variables. We extracted the average absolute temperature (AbsTemp) for each NUTS region at months m and m-1, and during the previous spring (March-May) seasons from a previously published dataset ^46^. We computed anomalies in temperature (AnomTemp) and in the Modified Normalized Difference Water Index (AnomMNDWI), as compared to the 2002-2024 period, at the NUTS level and at the same time steps by month (Table 2).

**Table 2.** Predictor variables used in the spatio-temporal occupancy model. Some variables were included as part of the “Circulation” (or presence) component of the occupancy model, while others as part of its “Reporting” (or detection) component.

| Variable | Description | Model component | Reference |
| --- | --- | --- | --- |
| **AbsTemp<sup>t</sup>** | Absolute temperature on period t | Circulation | 46 |
| **AnomTemp<sup>t</sup>** | Anomaly in temperature on period t | Circulation | 8 |
| **AnomMNDWI<sup>t</sup>** | Anomaly in MNDWI on period t | Circulation | 8 |
| **BRI** | Bird Risk Index averaged over the region | Circulation | 18 |
| **LogHumPop** | Log of the human population | Reporting | 48 |
| **Perc65+** | Proportion of the population aged 65 years old or more | Reporting | 48 |
| **GDPpc** | Gross Domestic Product per capita at country level | Reporting | 48 |
| **UrbClass** | Eurostat urban-rural typology (predominantly urban vs. others) | Reporting | 48 |
| **LogEqPop** | Log of the equine population | Reporting | See Supplementary Table S2 |
| **CountryUnderReport** | Countries that may under-report WNV equine cases as compared to human cases | Reporting | See Supplementary Figure S6 |
| **CountryOverReport** | Countries that may over-report WNV equine cases as compared to human cases | Reporting | See Supplementary Figure S6 |

Second, a fixed (structural) variable quantified the capacity of local ecosystems to sustain WNV reservoir circulation. Named the Bird Risk Index (BRI), it was computed at the NUTS region scale in a previously published study ^18^ by combining wild bird species spatial distribution and WNV serosurveys data in these species. We chose not to include any structural variable related to vectors, because *Cx. pipiens* is ubiquitous across Europe, hence heterogeneity in its abundance may not be considered structural but rather dependent on variations in weather conditions, i.e. on conjectural variables. What is more, published *Cx. pipiens* abundance maps (e.g. ^47^) themselves rely on climatic data.

Third, demographic and socio-economic variables were extracted from the Eurostat website ^48^ and prepared as described in the Supplementary Method 1. These NUTS-level variables were the log human population (LogHumPop), the proportion of the population aged 65 years old or more (Perc65+), the national Gross Domestic Product per capita (GDPpc), and the urban-rural typology (UrbClass) (Table 2). The latter was valued 1 if the NUTS was classified by Eurostat as predominantly urban, i.e. if at least 80% of the population lived in urban clusters, and 0 otherwise.

Fourth, we computed the log equine population in NUTS regions (LogEqPop). In absence of reliable standardized equine statistics in European regions, we retrieved estimates of national equine population sizes from gray literature sources in English or local languages (Supplementary Table S2). Moreover, the Grided Livestock of the World (GLW) database provides spatial distributions for several livestock species based on census statistics combined with modelling analyses ^49^. Because national equine population estimates we retrieved from this dataset were often low as compared to gray literature national estimates, we combined the latter with GLW within-country horse spatial distribution to compute the equine population by NUTS region.

Fifth, we identified countries that may under-report (CountryUnderReport) or over-report (CountryOverReport) WNV equine cases, as compared to WNV human cases. Indeed, we can assume the country-level WNV incidence rate to be similar in humans *vs*. in equids. Therefore, we identified countries that may under- and over-report equine cases as the points falling out of the prediction interval of a country-level linear model explaining the (population-weighted) number of NUTS-months with equine cases, with a 0 intercept and an offset of the (population-weighted) number of NUTS-months with human cases (see details in Supplementary Figure S6 and its legend). Germany was classified in the “CountryUnderReport” category, and Hungary and Romania in the “CountryOverReport” category.

### Spatio-temporal occupancy model

We implemented an occupancy model to predict the reporting (yes/no) of WNV human or equine cases in each NUTS region of the 27 European Union, for each month between June and October of 2010 to 2024. As already described in the literature ^38,39^, an occupancy model combines two components that quantify (i) the probability of true (unobserved) presence of a species, here the active circulation of WNV between birds and mosquitoes for a given NUTS region and month (the “Circulation” component, variable *z* below), and (ii) the probability of detection, here the reporting of human or equine cases caused by a bite from an infected mosquito (the “Reporting” component, variable *y* below) (Figure 1). We considered two models, respectively the “Human” and “Equine” models, depending on whether we used the human or equine cases data in the “Reporting” component. Even though the “Circulation” component had the same structure in both models, we did not combine both species in a single model. The reason was that we considered the infectious risk might be spatially heterogenous within NUTS regions and, because human and equine populations are not similarly distributed within regions, their exposures to WNV episytem might then differ. We scaled and centered all quantitative variables.

In the “Circulation” component, we determined explanatory variables (*X*^*circu*^ below) based on a DAG depicted in Supplementary Figure S1 where causal relationships were assumed between variables based on the literature (Supplementary Table S3) ^50^. Adjustment variables were, as described above, AnomTemp^m^, AnomTemp^m-1^, AnomTemp^prev spring^, AbsTemp^m^, AbsTemp^m-1^, AbsTemp^prev spring^, AnomMNDWI^m^, AnomMNDWI^m-1^, AnomMNDWI^prev spring^ and the BRI. Under the DAG assumptions, we were able to estimate a causal effect of some of the “Circulation” variables on the probability of WNV circulation in the bird reservoir, including conjectural (AnomTemp^m^ and AnomMNDWI^m^) and structural (BRI) variables. On the other hand, the coefficients of the other “Circulation” variables could only be interpreted as associations given the model’s adjustment set.

Predictors included in the “Reporting” component (*X*^*report*^ below) were demographic and socio-economic variables, namely LogHumPop, Perc65+, UrbClass and GDPpc in the “Human” model, and LogEqPop, CountryUnderReport and CountryOverReport in the “Equine” model (see Table 2 and hypothesized effects on WNV reporting in Supplementary Table S4).

Furthermore, WNV is more likely to spread in areas that were previously affected and to geographically close regions. To account for this spatial and temporal autocorrelation, we added both a Besag-York-Mollie 2 (BYM2) and a Random walk process at the year scale to the model. We fitted the model to the data using INLA ^39^.

Formally, if *z*_*i,t*_ and *y*_*i,t*_ are respectively the (latent) viral circulation and the (observed) reporting of cases in NUTS region *i* and month *t*:

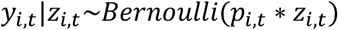

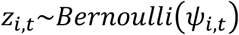

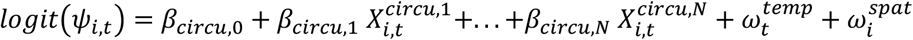

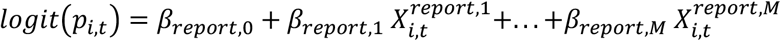

where *X*^*circu*^ and *X*^*report*^ are respectively the N “Circulation” component variables and the M “Reporting” component variables, 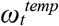 is the year-level Random walk process, and 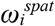 is the BYM2 process.

### Model validation

We first performed a cross-validation of the occupancy models by splitting the data into nine spatiotemporal blocks depicted in Supplementary Figure S7, as recommended by ^17,51^. For each model, we computed the pooled deviance-based marginal R^2^, as defined by ^52^, to check the consistency between model predictions and the testing data, as compared to a null model with the random effects only (including the spatial and temporal effects). An R^2^ value above 0 could then be interpreted as the full model predicting the testing data better than the null model. Then, in the final models, we computed areas under the ROC curve (AUC) using (i) the (human or equine) cases data used for model fitting and (ii) wild bird WNV outbreak data as reported to WAHIS, not used for model fitting.

### Sensitivity analysis

In a correctly specified DAG, multiple adjustment sets should approximately yield to the same causal effect estimates. To assess the sensitivity of our results to the adjustment variables included in the DAG, we fitted subsets of the model by removing one or several of the variables that should not, if the DAG is true, add substantial bias to the causal effect estimate (see details in Supplementary Table S5).

### Attributable fractions

Within the model’s “Circulation” component, we computed population attributable fractions (PAF) for the structural variable on the one hand, and for the group of conjectural variables on the other hand. We accounted for the variables’ causal structure and continuous nature, based on previously published methods ^19–21^. For a given variable *X*^*circu,k*^, the PAF was calculated across NUTS regions *i* and months *t*:

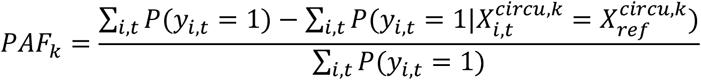

The so-called “intervention” was thus to set 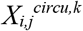 values to 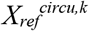, which was the median value of the lowest (in case of positive effect of the variable) or highest (in case of negative effect) decile of *X*^*circu,k*^.

The structural PAF was composed of the BRI only, but the conjectural PAF was computed from all weather variables with a previously described sequential method ^20,21^. Briefly, a sequence of conjectural variables was randomly ordered. The above intervention was applied to the first variable of the sequence, and we accounted for its effect on the whole dataset, i.e. on the outcome *y* but also on the other explanatory variables assumed to be descendants of the first variable in the DAG. On the next step, the same was applied to the second variable, then the third, etc. The PAF of the overall sequence was the sum of all these individual PAF. The process was repeated for 6 different sequences and the final conjectural PAF was the average of these repetitions. This method was applied both globally and for each NUTS-month.

## Data Availability

The code reproducing this study is available at: https://github.com/JonathanBas/WNV_Europe.

## Acknowledgements

We acknowledge the European Centre for Disease Prevention and Control for providing the human cases data, and Pierre-Yves Boëlle for useful comments. This work was supported by a DIM1Health postdoctoral fellowship awarded by the Conseil Régional d’Ile-de-France, and by a French State funding managed by the National Research Agency under France 2030, as part of the PEPR PREZODE project (reference ANR 24 PEPZ 0004). The funders had no role in study design, data collection and analysis, decision to publish or preparation of the manuscript.

