## Supplementary information for "Short- and long-term causes of West Nile virus risk in Europe: a spatiotemporal model accounting for under-reporting"

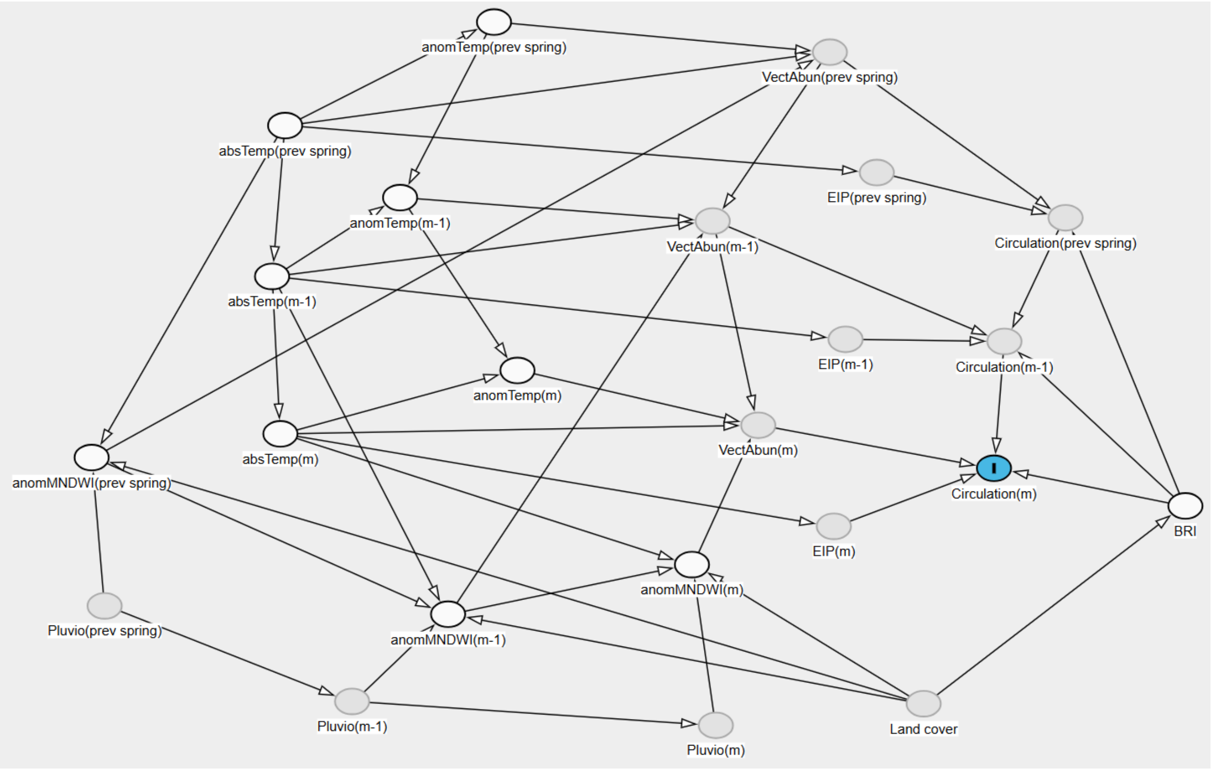


**Supplementary Figure S1.** Directed Acyclic Graph (DAG) of the “Circulation” component of the spatio-temporal occupancy model. Variables displayed in white are included in the model, while variables displayed in gray are unobserved. Based on this DAG, we can estimate the causal effect on the outcome (circulation on month m, displayed here in blue) of the BRI, the MNDWI anomaly on month m, and temperature anomaly on month m. Here, we suppose that the paths going through m-k (with $k \geq 2$) induce only weak biases, hence we do not account for them. Abbreviations: m = month between June and October; BRI = Bird Risk Index; absTemp = absolute temperature; anomTemp = temperature anomaly; anomMNDWI = MNDWI anomaly; Pluvio = pluviometry; EIP = WNV extrinsic incubation period; VectAbun = *Culex* vector abundance. Figure created with Dagitty.net.

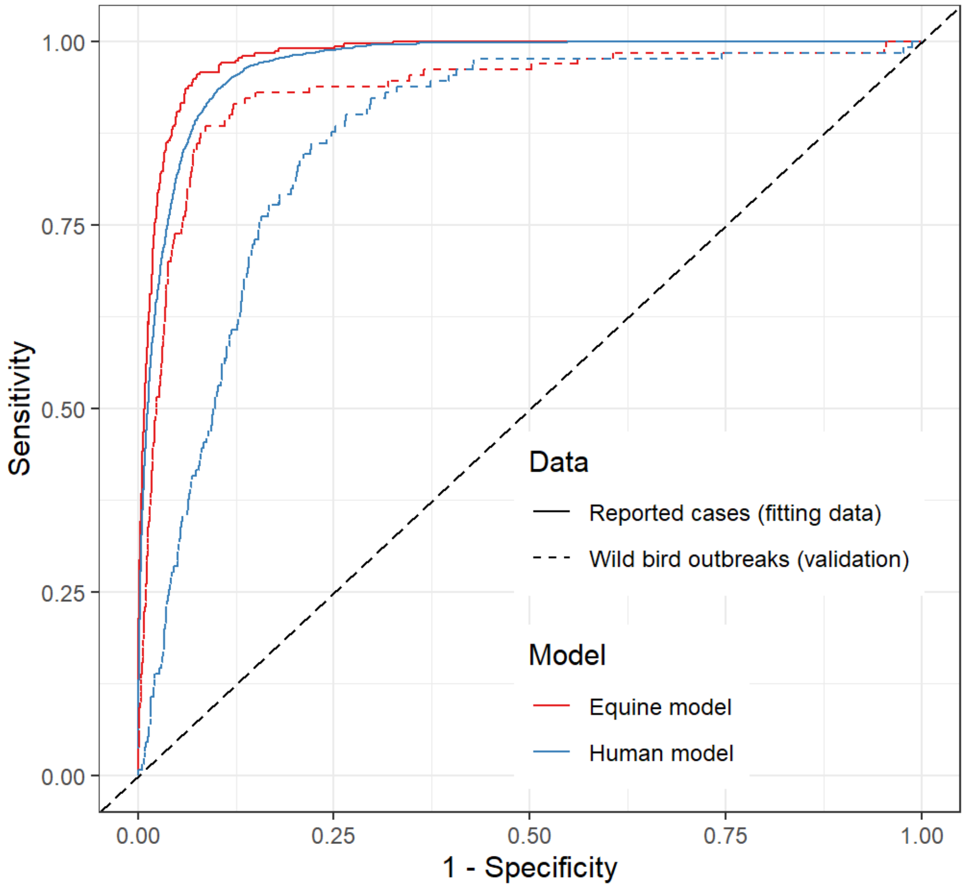


**Supplementary Figure S2.** Receiver operating characteristic (ROC) curves for the "Equine" and "Human" model predictions, as compared to the reported (equine or human) cases (data used for model fitting) and to the wild bird outbreaks notified to WAHIS (data used for validation).

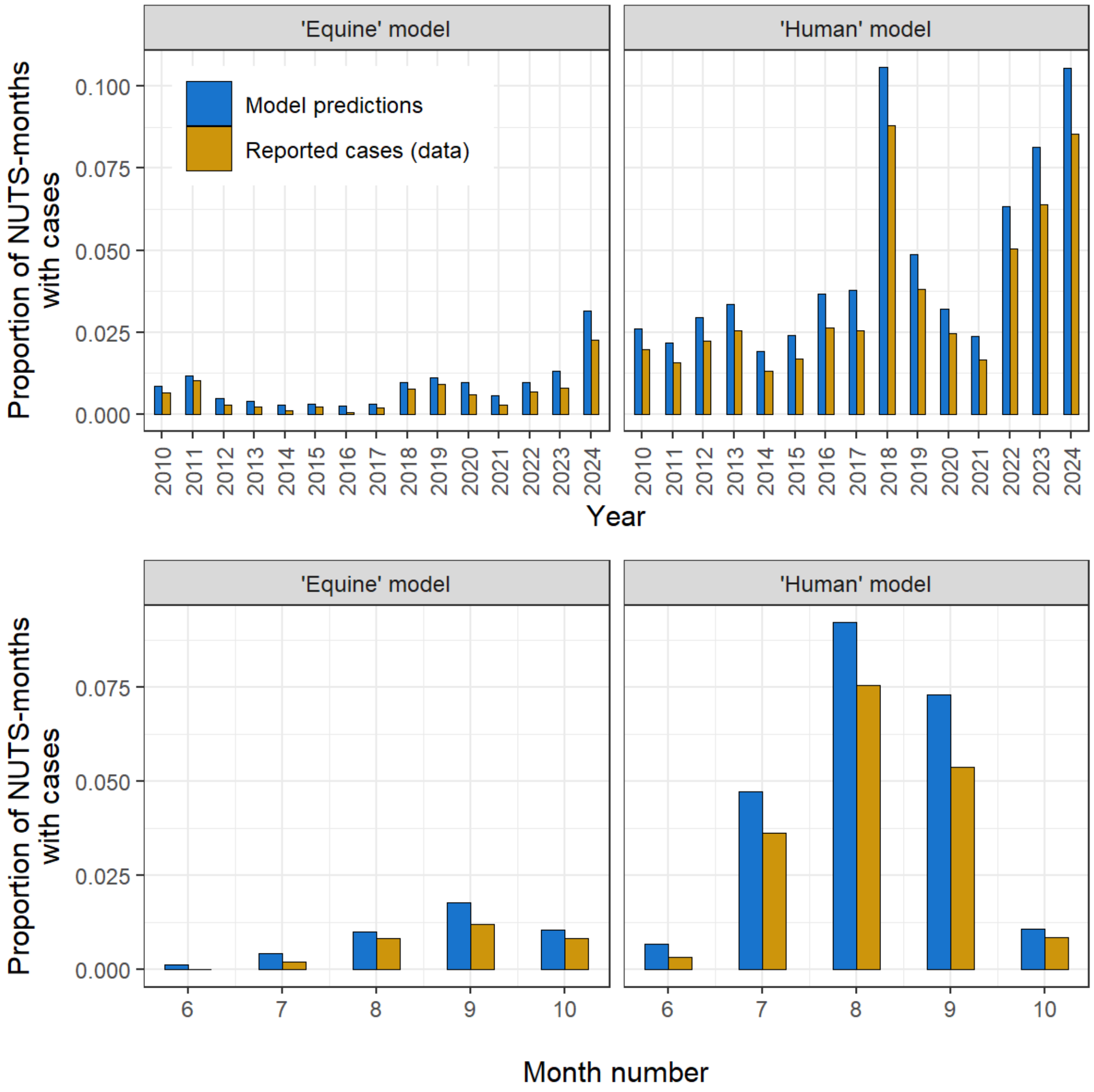


**Supplementary Figure S3.** Proportion of NUTS-months with reported cases, as observed in the data and as predicted by the "Equine" (left panels) and "Human" (right panels) models, by year (top panels) and by month of the year (bottom panels).

**Supplementary Table S1. Odds-ratios of the "Circulation" component variables.** The models were fitted to a dataset where all variables were scaled. In this table, we back-transform the estimated coefficient values in order to display adjusted odds-ratio (aOR) values, and their 95% credible interval, relatively to changes of unscaled explanatory variables (original data). Effects that are different from 1 and that may be interpreted, under our DAG assumptions, as causal effects are displayed in bold.

| **Variable** | **Type of coefficient** | **Increase of the explanatory variable** | **"Equine" model aOR** | **"Human" model aOR** |
|---|---|---|---|---|
| BRI | Causal effect | +1 interquartile range | 1.08 [0.62; 1.87] | **1.85 [1.30; 3.44]** |
| $AnomMNDWI^{m}$ | Causal effect | +1 standard deviation as compared to 2002-2024 period | 0.98 [0.57; 1.67] | 1.05 [0.82; 1.33] |
| $AnomTemp^{m}$ | Causal effect | +1 standard deviation as compared to 2002-2024 period | **1.61 [1.28; 2.32]** | 1.03 [0.95; 1.12] |
| $AnomMNDWI^{m-1}$ | Association# | +1 standard deviation as compared to 2002-2024 period | 0.38 [0.17; 0.71] | 0.94 [0.73; 1.19] |
| $AnomMNDWI^{spring}$ | Association# | +1 standard deviation as compared to 2002-2024 period | 3.45 [1.58; 9.65] | 2.45 [1.7; 4.06] |
| $AnomTemp^{m-1}$ | Association# | +1 standard deviation as compared to 2002-2024 period | 0.59 [0.4; 0.75] | 0.52 [0.37; 0.59] |
| $AnomTemp^{spring}$ | Association# | +1 standard deviation as compared to 2002-2024 period | 0.74 [0.48; 1.13] | 1.75 [1.4; 3.13] |
| $AbsTemp^{m}$ | Association# | +1°C | 0.74 [0.61; 0.81] | 1.09 [1.07; 1.14] |
| $AbsTemp^{m-1}$ | Association# | +1°C | 1.79 [1.53; 2.65] | 1.76 [1.64; 2.27] |
| $AbsTemp^{spring}$ | Association# | +1°C | 1.68 [1.30; 2.24] | 0.93 [0.76; 1.03] |

aOR: adjusted odds-ratio.

### Association between the explanatory variable and the outcome, given the model's adjustment set, which cannot be considered as causal effects under our DAG assumptions.

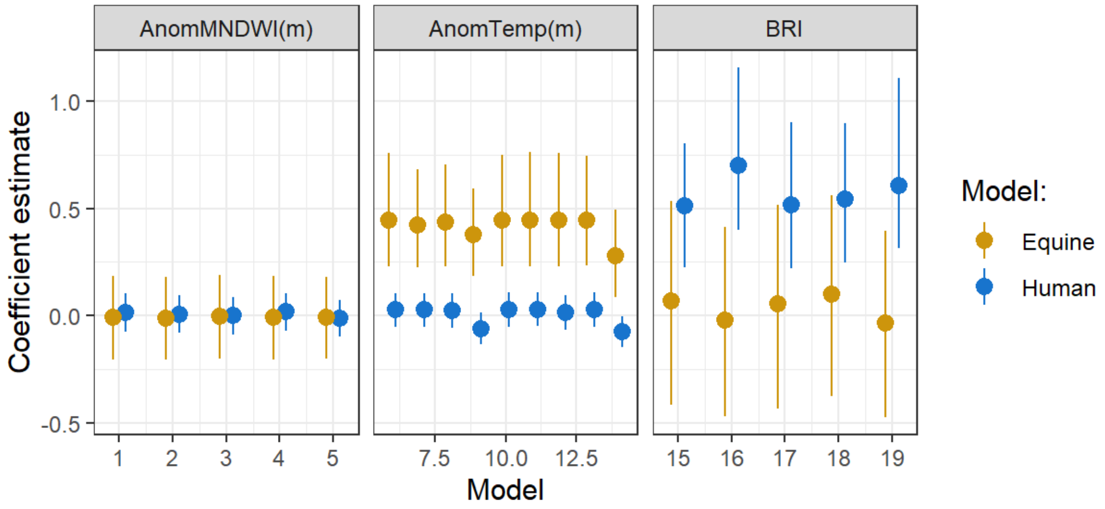


**Supplementary Figure S4.** Results of the sensitivity analysis. Both "Human" and "Equine" models were fitted to the data while considering different adjustment sets to infer causal effects for three variables (adjustment sets detailed in Supplementary Table S5).

**Supplementary Method S1.** Data preparation.

Equine cases data. The World Animal Health Information System (WAHIS) website includes two types of data on WNV equine cases:

- the "Events" data with reports including information on the species affected, time and GPS coordinates of WNV outbreaks in animals
- the "Semestrial" data with WNV animal cases reported at the semester scale (January-June and July-December) and at various geographical scales (from the NUTS3 region to the country).

Both datasets are not always consistent with one another. Because of scale discrepancies, we were only able to integrate the "Events" data to the model, at the NUTS and month scales, although the "Semestrial" data sometimes suggested that WNV cases may be occurring without being reported in the "Events" data. For instance, one or several WNV equines cases were sometimes reported in the "Semestrial" data in country A during the January-June semester of year X, but without report in the "Events" data. In that case, we considered that equine case observations in all NUTS regions of country A and all months of the January-June semester of year X were NAs.

*Socio-economic data*. It was obtained from Eurostat website [1] as of October 2025. Variables were the log human population by NUTS and year (LogHumPop), the proportion of the population aged 65 years old or more by NUTS and year (Perc65+), the Gross Domestic Product per capita by country and year (GDPpc), the percentage of GDP spent in Health (PercHealthGDP) and in Agriculture (PercAgriGDP) by country and year, and the urban-rural typology for each NUTS (UrbClass). Data for some years and regions was missing. If the GDPpc or Perc65+ was missing for a NUTS3 region (respectively for a NUTS2 region), we then used the value of the larger NUTS2 region (resp. NUTS1 region). Values of LogHumPop between 2010 and 2013 were missing and were assigned the 2014 value. The 2024 value of GDPpc was missing for many regions, and was in that case assigned the 2023 value. Finally, Perc65+ was missing for NUTS regions of Croatia in 2013, and were therefore assigned their 2012 value.

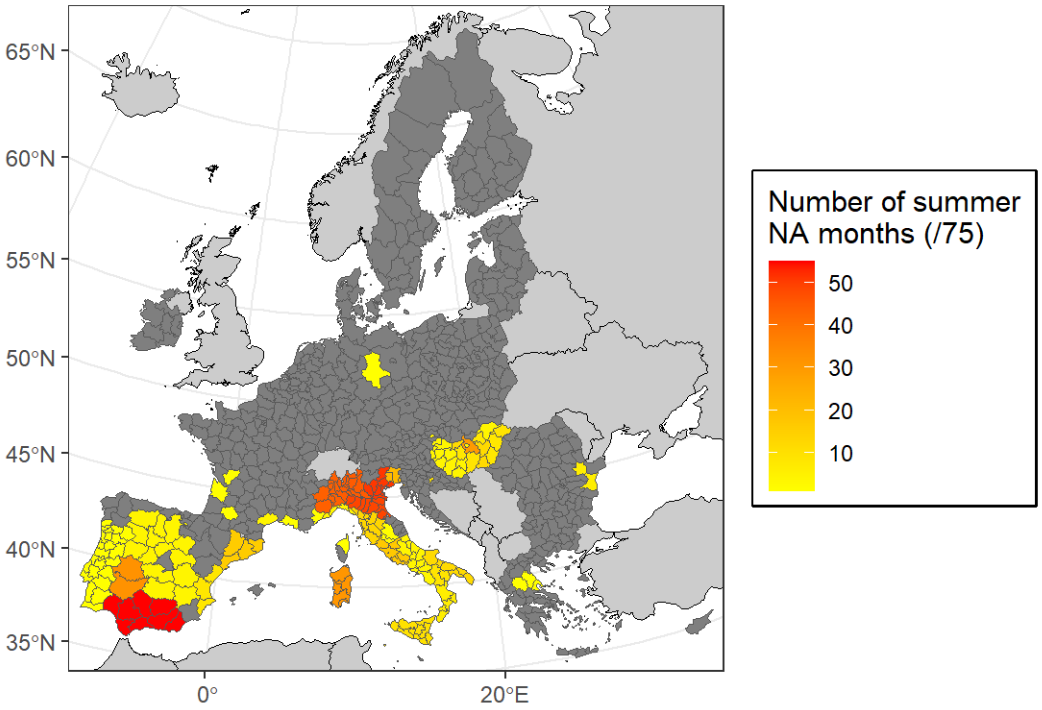


**Supplementary Figure S5.** Number of months with missing information (NA) on the reporting of WNV equine cases by NUTS region, across the study period. The total number of months included in the study is 75 (June to October between 2010 and 2024). NUTS with zero month with missing information appear in dark gray.

**Supplementary Table S2.** Gray literature sources for equine population estimates in European countries.

| Country | Country code | Total equine population | Source |
|---|---|---|---|
| Austria | AT | 140,000 | https://www.bmluk.gv.at/themen/landwirtschaft/landwirtschaft-in-oesterreich/tierische-produktion/pferde/tier_prod_pferde.html |
| Belgium | BE | 397,140 | https://montlesoie.be/storage/documents/cNllammreq3unyebxQL4yGLWtsV4Fayv3hT95opr.pdf |
| Bulgaria | BG | 121,649 | WorldHorseWelfare and Eurogroups for Animals - Removing the Blinkers: The Health and Welfare of European Equidae in 2015 |
| Cyprus | CY | 7,350 | WorldHorseWelfare and Eurogroups for Animals - Removing the Blinkers: The Health and Welfare of European Equidae in 2015 |
| Czech Republic | CZ | 106,643 | https://www.jezdci.cz/clanky/za-rok-pribylo-v-ceske-republice-tri-tisice-koni-je-jich-temer-105-tisic/ |
| Germany | DE | 461,000 | WorldHorseWelfare and Eurogroups for Animals - Removing the Blinkers: The Health and Welfare of European Equidae in 2015 |
| Denmark | DK | 180,000 | https://www.ridehesten.com/nyheder/en-lille-historie-om-hesten-i-danmark/74389 |
| Estonia | EE | 14,864 | https://www.pria.ee/registrid/avalikud-andmed#loomade-statistilised-andmed-hetkeseisuga |
| Greece | EL | 70,443 | WorldHorseWelfare and Eurogroups for Animals - Removing the Blinkers: The Health and Welfare of European Equidae in 2015 |
| Spain | ES | 638,979 | EL SECTOR EQUINO ESPAÑOL EN 2024: PRINCIPALES MAGNITUDES E INDICADORES ECONÓMICOS |
| Finland | FI | 72,000 | HEVOSTALOUS LUKUINA 2022 (https://www.ratsastus.fi/site/assets/files/28620/hevostalous_lukuina_2022_lopullinen.pdf) |
| France | FR | 1,005,200 | IFCE CHIFFRES CLÉS 2024 |
| Croatia | HR | 43,400 | https://gospodarski.hr/rubrike/stocarstvo-rubrike/pozitivan-trend-razvitka-konjogojstva-u-hrvatskoj/#:~:text=Brojno%20stanje%20konja%20u%20Hrvatskoj,36.000%20konja%20i%207.400%20magaraca. |
| Hungary | HU | 43,200 | KSH (https://www.ksh.hu/stadat_files/mez/hu/mez0027.html) |
| Ireland | IE | 120,912 | Equine census report 2024-25 (https://assets.gov.ie/static/documents/7b4997a3/Census_report_Nov_2025_final.pdf) |
| Italy | IT | 450,000 | senato.it/japp/bgt/showdoc/REST/v1/showdoc/get/fragment/18/DDLPRES/0/1300099/all |
| Lithuania | LT | 12,500 | OSP (https://osp.stat.gov.lt/lietuvos-aplinka-zemes-ukis-ir-energetika-2022/zemes-ukis/gyvulininkyste) |
| Luxembourg | LU | 3,600 | https://paperjam.lu/article/news-lequitation-une-passion-un-secteur-economique |

| | | | |
|---|---|---|---|
| Latvia | LV | 17,696 | OSP (https://data.stat.gov.lv/pxweb/lv/OSP_OD/OSP_OD__skait_apsek__dzivnieki__laukskait/LSK01-III47.px/) |
| Malta | MT | 1,860 | WorldHorseWelfare and Eurogroups for Animals - Removing the Blinkers: The Health and Welfare of European Equidae in 2015 |
| Netherlands | NL | 450,000 | NEDERLAND PAARDENLAND FEITEN & CIJFERS (https://www.kennisbanksportenbewegen.nl/?file=7063&m=1467808613&action=file.download) |
| Poland | PL | 309,964 | Aktualny stan hodowli koni w Polsce, Patrycja Wojciechowska, Ewa Metera-Zarzycka (https://www.kpodr.pl/wp-content/uploads/2024/12/Aktualny-stan-hodowli-koni-w-Polsce.pdf) |
| Portugal | PT | 99,948 | Anuário do Cavalo, Horse Economic Forum (https://asset.skoiy.com/a6e66712e23e7657db1c9e8aa6c4d037/6jhyahh4nszx.pdf) |
| Romania | RO | 744,438 | WorldHorseWelfare and Eurogroups for Animals - Removing the Blinkers: The Health and Welfare of European Equidae in 2015 |
| Sweden | SE | 355,000 | https://hastnaringen-i-siffror.se/ |
| Slovenia | SI | 18,043 | SiStat (https://pxweb.stat.si/SiStatData/pxweb/sl/Data/-/1516606S.PX) |
| Slovakia | SK | 22,550 | https://hnonline.sk/slovensko/24295589-pocet-koni-je-u-nas-najvyssi-za-40-rokov-zasadne-sa-zmenilo-aj-ich-vyuzitie |

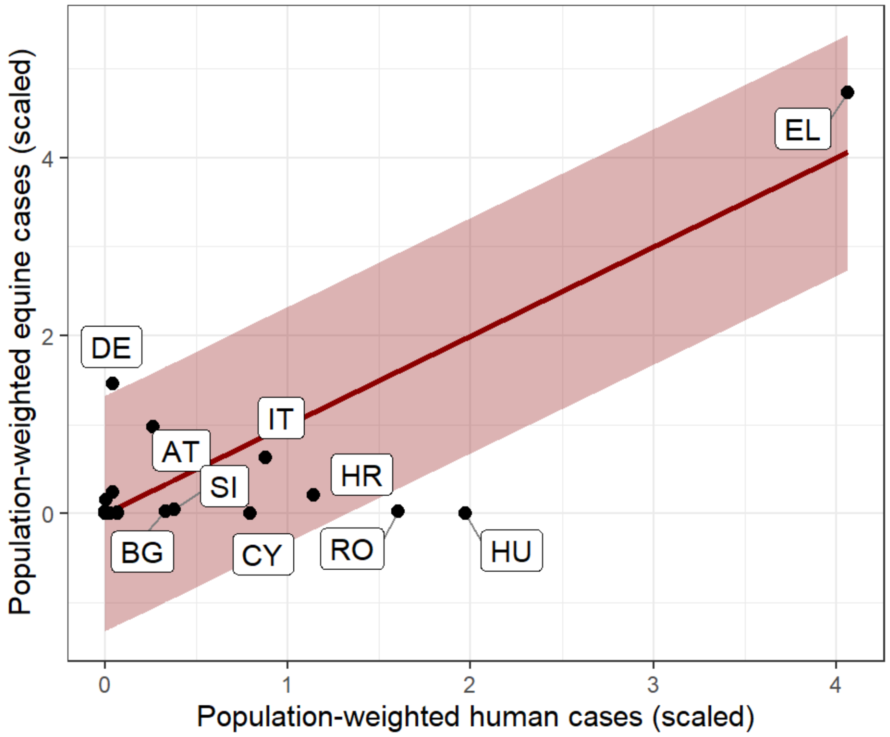


**Supplementary Figure S6.** Relationship between the cumulated number of NUTS-months with notified WNV human and equine cases in European countries, between 2010 and 2024, weighted by the human and equine populations and scaled. The red line and prediction interval are that of a linear regression explaining the second variable, with a 0 intercept and an offset of the first variable. Countries below (respectively above) the prediction interval are considered to potentially under-report (resp. over-report) WNV equine cases as compared to WNV human cases.

**Supplementary Table S3.** Causal relationships between variables of the "Circulation" model's component, as assumed in the Directed Acyclic Graph (DAG).

| Causal relationship | References or rationale |
|---|---|
| Effect of the temperature and pluviometry on month m-1 on the temperature and pluviometry on month m | [2,3] |
| Effect of absolute temperature on temperature anomaly | By definition |
| Effect of land cover on MNDWI | [4] |
| Effect of temperature on the water index | [5] |
| Effect of absolute temperature on month m on *Culex* mosquito abundance on month m [(a)] | [6–8] |
| Effect of temperature anomaly on month m on *Culex* mosquito abundance on month m [(a)] | [9–12] |
| Effect of the water index anomaly on month m on *Culex* mosquito abundance on month m | [12,13] |
| Effect of mosquito vector abundance on the circulation of WNV | [14] |
| Effect of temperature on WNV extrinsic incubation period (EIP) | [7,8] |
| Effect of WNV extrinsic incubation period (EIP) on the circulation of WNV | [7,8] |
| Effect of land cover on the Bird Risk Index (BRI) | By construction ([15]) |
| Effect of the Bird Risk Index (BRI) on the circulation of WNV | [15] |

[(a)] We considered that field studies allowed to evaluate the effect of temperature **anomaly** on mosquito abundance when they compared different time periods within the same global geographical area, while laboratory studies allowed to evaluate the effect of **absolute** temperature.

**Supplementary Table S4.** Hypothesized relationships between variables of the “Reporting” model’s component and WNV (human and equine) case reporting.

| Model | Variable | Hypothesis | Expected relationship |
|---|---|---|---|
| **“Human” model** | Log of the human population (LogHumPop) | More humans susceptible to be infected by WNV. | Positive |
| | Proportion of the population aged 65 years old or more (Perc65+) | Older individuals are more likely to present severe forms of WNV infection, and thus to be hospitalized and notified to the surveillance system. | Positive |
| | Gross Domestic Product per capita (GDPpc) | A higher GDP may lead to a more performant public health surveillance system, and thus a higher reporting fraction of cases. | Positive |
| | Urban-rural typology (predominantly urban vs. others) (UrbClass) | The urban *vs.* rural habitat may affect human exposure to *Culex* mosquitoes that previously bit infectious birds. | Negative |
| **“Equine” model** | Log of the equine population (LogEqPop) | More equids susceptible to be infected by WNV. | Positive |
| | Countries that may under-report WNV equine cases as compared to human cases (CountryUnderReport) | By definition, these countries may under-report WNV equine cases. | Negative |
| | Countries that may over-report WNV equine cases as compared to human cases (CountryOverReport) | By definition, these countries may over-report WNV equine cases. | Positive |

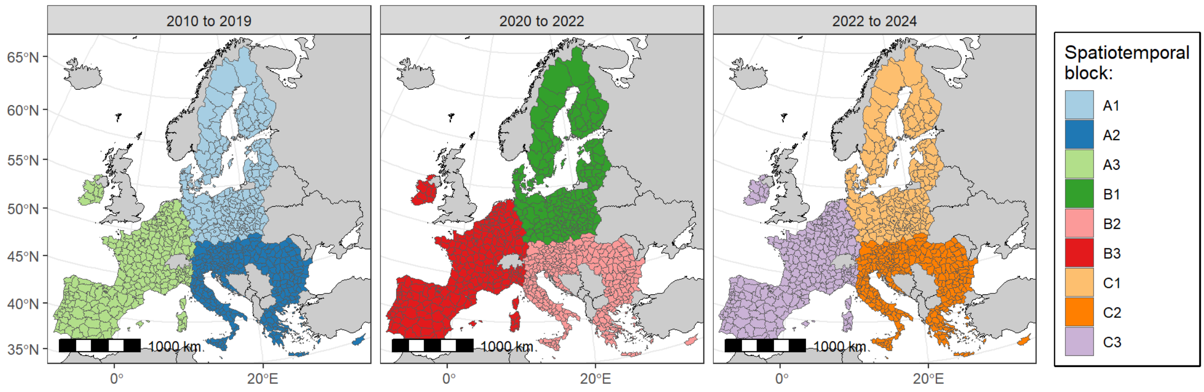


**Supplementary Figure S7.** Spatiotemporal blocks used for the nine-fold cross-validation of the model. Time splits are 2010-06 to 2019-10 (blocks A1, A2 and A3), 2020-06 to 2022-06 (blocks B1, B2 and B3), and 2022-07 to 2024-10 (blocks C1, C2 and C3).

**Supplementary Table S5.** Models fitted as part of the sensitivity analysis. Subsets of the main analysis (Human and Equine) models were fitted to the same dataset, to check whether different adjustment sets in the "Circulation" model component lead to the same causal effect estimates for target variables.

| Model | Causal effect assessed | Adjustment set |
|---|---|---|
| 1 | AnomMNDWI$^{m}$ | All main analysis variables |
| 2 | AnomMNDWI$^{m}$ | All main analysis variables excepted AnomTemp$^{\text{prev spring}}$ |
| 3 | AnomMNDWI$^{m}$ | All main analysis variables excepted AnomTemp$^{m-1}$ |
| 4 | AnomMNDWI$^{m}$ | All main analysis variables excepted AnomTemp$^{m}$ |
| 5 | AnomMNDWI$^{m}$ | All main analysis variables excepted AnomTemp$^{\text{prev spring}}$, AnomTemp$^{m-1}$ and AnomTemp$^{m}$ |
| 6 | AnomTemp$^{m}$ | All main analysis variables |
| 7 | AnomTemp$^{m}$ | All main analysis variables excepted AbsTemp$^{\text{prev spring}}$ |
| 8 | AnomTemp$^{m}$ | All main analysis variables excepted AnomTemp$^{\text{prev spring}}$ |
| 9 | AnomTemp$^{m}$ | All main analysis variables excepted AbsTemp$^{m-1}$ |
| 10 | AnomTemp$^{m}$ | All main analysis variables excepted AnomMNDWI$^{m}$ |
| 11 | AnomTemp$^{m}$ | All main analysis variables excepted AnomMNDWI$^{m-1}$ |
| 12 | AnomTemp$^{m}$ | All main analysis variables excepted AnomMNDWI$^{\text{prev spring}}$ |
| 13 | AnomTemp$^{m}$ | All main analysis variables excepted BRI |
| 14 | AnomTemp$^{m}$ | AnomTemp$^{m}$, AnomTemp$^{m-1}$ and AbsTemp$^{m}$ |
| 15 | BRI | All main analysis variables |
| 16 | BRI | All main analysis variables excepted AnomTemp$^{\text{prev spring}}$ |
| 17 | BRI | All main analysis variables excepted AnomTemp$^{m-1}$ |
| 18 | BRI | All main analysis variables excepted AnomTemp$^{m}$ |
| 19 | BRI | All main analysis variables excepted AnomTemp$^{\text{prev spring}}$, AnomTemp$^{m-1}$ and AnomTemp$^{m}$ |
